# Comparison of metabolic syndrome prevalence using three different definitions: A population-based study in Peru

**DOI:** 10.1101/2023.10.23.23297432

**Authors:** Víctor Juan Vera-Ponce, Fiorella E. Zuzunaga-Montoya, Luisa Erika Milagros Vásquez Romero, Joan A. Loayza-Castro, Mario J. Valladares-Garrido

**Affiliations:** Instituto de Investigación en Ciencias Biomédicas de la Universidad Ricardo Palma; Universidad Tecnológica del Perú, Lima, Peru; Universidad Científica del Sur, Lima, Perú; Universidad Continental Lima, Perú; Oficina de Epidemiología, Hospital Regional Lambayeque, Chiclayo. Peru

**Keywords:** Metabolic syndrome, epidemiologic factors, public health (source: MeSH NLM)

## Abstract

**Introduction:** Metabolic syndrome (MetS) is important for public health; however, there are different ways to define it.

**Objectives:** 1) to estimate the prevalence of MetS using three different definitions: the criteria of the International Diabetes Federation (IDF), the World Health Organization (WHO), and the National Cholesterol Education Program’s Adult Treatment Panel III (ATPIII), 2) to identify the factors associated with the presence of MetS according to each criterion, and 3) to evaluate the agreement between these three.

**Materials and Methods:** A secondary and cross-sectional analysis of the database from the Life Stages Food and Nutrition Surveillance Survey (VIANEV) was conducted. For the definition of MetS, the aforementioned definitions were used. A multivariable Poisson regression analysis with robust variance and agreement was evaluated through the Kappa index.

**Results:** According to ATPIII, IDF, and WHO, the prevalence of MetS was 42.60%, 46.78%, and 49.49%, respectively. The agreement between IDF with WHO and ATPIII criteria was 0.42 and 0.45, while for ATPIII and WHO it was 0.44. In general, the associated factors were identified as sex, marital status, age, region of residence, level of physical activity, smoking habit, and body mass index (BMI). However, the association of these varied according to the definition used.

**Conclusion:** The prevalence of MetS varies significantly according to the criteria used. This was higher when the WHO definition was used compared to the others. Additionally, the associated factors varied according to the definition used, although a consistency was observed across all definitions with BMI.

## Introduction

Metabolic syndrome (MetS) is a set of conditions that, when occurring together, significantly increase the risk of stroke, heart disease, and type 2 diabetes ^(1,2)^. The consequences of MetS are profound, as it can lead to increased mortality, a decrease in quality of life, and a significant economic burden on health systems ^(3)^.

Data from the 2009 National Health and Nutrition Examination Survey suggested that nearly one in three (34.7%) American adults during that period were impacted by Metabolic Syndrome or dealing with its assorted symptoms and related health issues ^(4)^. A 2011 systematic review analyzing Latin American studies found an average of almost a quarter of individuals (24.9%) afflicted (6). Yet it’s crucial to note that prevalence varies greatly between nations in the region, like Peru, relying on the demographics examined and standards followed; some research places the number anywhere from one-fifth to nearly half ^(7–9)^.

The diagnosis of MetS presented issues due to various criteria existing. Commonly used were those of the Adult Treatment Panel III (ATPIII), the International Diabetes Federation (IDF), and the World Health Organization (WHO) ^(10)^. However, despite focusing on mutual metabolic risks, the thresholds and risks delineated diverged significantly. Distinctions in criteria led to prevalence differing reliant on what was utilized ^(11–13)^. This proposed the sets failed to encapsulate alike people with MetS fully. This wavering prevalence depends on standards highlighted requiring greater harmony and lucidity in the condition’s definition.

This research tackles some uncertainties surrounding MetS due to its significance as a public health issue and doubts about its identification and related aspects. It aims to handle a few of these unknowns. To start, it intended to ascertain the occurrence of MetS in a population of Peru relying on diagnostic standards of ATPIII, IDF, and WHO. Second, it wanted to evaluate the contract between these 3 sets of standards. Since all are intended to pinpoint the identical underlying condition, a high contract between them is anticipated. Finally, it seeks to recognize the factors linked to MetS in our study group and decide if they are uniform among the different sets of criteria. Given that all sets of criteria are presumed to assess metabolic syndrome, it’s expected that the associated factors will be almost identical among them.

Through these objectives, it is hoped to provide greater clarity on the prevalence and factors associated with MetS in Peru and contribute to the understanding of how different diagnostic criteria can influence these estimates.

## METHODS

### Study Design and Context

This research is a secondary and cross-sectional analysis of the database from the Life Stages Food and Nutrition Surveillance Survey (VIANEV), carried out during the 2017-2018 period, developed and administered by the National Center for Food and Nutrition (CENAN) of Peru ^(14)^. An analytical and concordance approach was used to determine the proposed objectives.

To ensure the quality and transparency of our work, we followed the STROBE (Strengthening the Reporting of Observational studies in Epidemiology) guidelines ^(15)^. These guidelines provide a framework for the proper presentation of observational research, ensuring that all important aspects of design, analysis methods, and findings are reported.

### Population, Sample, and Eligibility Criteria

Through VIANEV, data were collected from three distinct domains: Metropolitan Lima, the capital of Peru, and other urban and rural areas of the country. Data collection was done through a stratified, multistage, and probabilistic sampling process, which was independent in each domain. The sample selection was carried out in two stages: In the first, clusters of primary sampling units were randomly selected, and in the second, households were randomly selected within these clusters that included adults aged 18 to 59 years. Thanks to this sampling process, inferences could be made both at the national level and in urban, rural, and Metropolitan Lima areas. For more information on the VIANEV survey methodology, you can review the survey’s technical report and previous studies ^(14)^.

After considering specific exclusion criteria, in the present study, people who did not present the variables that make up MetS, such as those with a history of arterial hypertension or type 2 diabetes mellitus, were not considered.

### Definition of Variables

The main variable was MetS, whose definition was based on the three most common criteria. According to ATPIII, MetS is defined as the presence of three or more factors: abdominal obesity (waist circumference greater than 102 cm in males and 88 cm in females), elevated triglycerides (≥150 mg/dL), low HDL (<40 mg/dL in men and <50 mg/dL in women), high blood pressure (≥130/85 mmHg or treatment for hypertension), and elevated fasting glucose (≥110 mg/dL or treatment for hyperglycemia) ^(16)^.

The IDF defines MetS as the presence of abdominal obesity (waist circumference greater than 94 cm in men and 80 cm in women) plus two of the other four factors used by ATPIII ^(17)^.

The WHO, on the other hand, defines MetS as the presence of altered fasting glucose (≥ 110 mg/dl), plus two of the following: obesity (body mass index greater than 30 kg/m²), dyslipidemia (triglycerides ≥150 mg/dL or HDL <35 mg/dL in men and <39 mg/dL in women), high blood pressure (≥140/90 mmHg). It also considers microalbuminuria (urinary excretion of albumin ≥20 µg/min or albumin/creatinine ratio ≥30 mg/g), although it was not included ^(18)^.

The covariables considered in this research, as possible factors associated with MetS, include demographic, socioeconomic, behavioral, and health characteristics. Demographic traits such as sex, whether male or female; age group of 18 to 29 years, 30 to 39 years, 40 to 49 years, and 50 to 59 years old; marital status of single or partnered; and highest attained educational level of primary school, secondary school, or high school were the characteristics examined. Socioeconomic characteristics include the natural region (Metropolitan Lima, rest of the coast, highlands, and jungle), area of residence (urban, rural), and socioeconomic level (poor, not poor). Behavioral characteristics encompassed whether alcohol had been consumed within the last month and if the individual presently smoked, each factor limited to a binary yes or no response. Physical activity levels and body mass index, which characterize health, can be low or high as well as normal, overweight, or obese.

These factors were selected based on their theoretical and empirical relevance in the existing literature on MetS. The way each variable was measured can be reviewed in the VIANEV report ^(14)^.

### Statistical Analysis

Data analysis was carried out using R software version 4.0.5. Descriptive variables were presented in terms of absolute and relative frequencies. Associated factors were evaluated in a bivariate analysis, and the calculation of crude (RPc) and adjusted prevalence ratios (RPa) with their respective 95% confidence intervals (CI95%) was made. For this, generalized linear models with robust variance estimation were used, and a Poisson distribution with logarithmic link functions was assumed. In addition, concordance analyses were carried out to evaluate the consistency between the different sets of diagnostic criteria for MetS. In addition, a bar graph was made to reflect the distribution of the components of MetS, according to the cut-off point established by each one.

Likewise, a concordance analysis was carried out with the purpose of evaluating the consistency between the different diagnostic criteria for MetS, using the Kappa coefficient, a statistical measure that quantifies the degree of agreement between two observers or measurement methods, beyond the agreement that would be expected by chance. A Kappa coefficient of 1 indicates perfect agreement, and one that any observed agreement is purely coincidental. In the results of this analysis, there is an additional view on the consistency between the three diagnostic criteria already indicated. To visualize the overlap and differences between the cases of MetS identified by each of the diagnostic criteria, a Venn diagram was also created.

All analyses were carried out according to the complexity of the sample design; that is, stratification, clustering, and sample weights were taken into account in all statistical calculations.

### Ethical Aspect

This article was developed using data sets from the VIANEV Survey that are freely available and in the public domain online (http://iinei.inei.gob.pe/microdatos/). All personal identifiers had been removed from these data sets before their publication, ensuring the anonymity of the patients.

Given that the work relied on an analysis of preexisting anonymous information collected elsewhere, submitting it for assessment by an ethics board was deemed unnecessary. This method aligns with ethical health research guidelines, specifying analyses of publicly accessible datasets devoid of identifiable details require no formal ethical review, though safeguards were instituted to ensure the analysis proceeded ethically and respectfully regarding participants’ rights and dignity in the original research.

## RESULTS

Table 1 contains the characteristics of the research sample: 885 individuals. In terms of sex, 55.59% were women (n=492). In terms of area of residence, 32.43% lived in rural areas (n=287). In relation to the prevalences of MetS according to the different diagnostic criteria of ATPIII, 42.60% had MetS (n=377); according to IDF criteria, 46.78% had MetS (n=414), while according to WHO criteria, 49.49% had MetS (n=438). The total prevalence of MetS is 66.78%.

**Table 1.** Demographic, socioeconomic, behavioral, and health characteristics of study participants and prevalence of metabolic syndrome.

| Characteristic | n = 885 |
| --- | --- |
| **Sex** |  |
| Female | 492.00 (55.59%) |
| Male | 393.00 (44.41%) |
| **Age group** |  |
| 18 to 29 years old | 246.00 (27.80%) |
| 30 to 39 years old | 223.00 (25.20%) |
| 40 to 49 years old | 215.00 (24.29%) |
| 50 to 59 years old | 201.00 (22.71%) |
| **Civil status** |  |
| Single | 318.00 (35.93%) |
| With couple | 567.00 (64.07%) |
| **Educational level** |  |
| Higher | 346.00 (39.27%) |
| Secondary | 336.00 (38.14%) |
| Until primary | 199.00 (22.59%) |
| **Natural region** |  |
| Jungle | 130.00 (14.69%) |
| Metropolitan Lima | 415.00 (46.89%) |
| Mountain Range | 157.00 (17.74%) |
| Resy of coast | 183.00 (20.68%) |
| **Area of residence** |  |
| Rural | 287.00 (32.43%) |
| Urban | 598.00 (67.57%) |
| **Wealth index** |  |
| No poor | 721.00 (81.47%) |
| Poor | 164.00 (18.53%) |
| **Alcohol consumption** |  |
| No | 450.00 (50.85%) |
| Yes | 435.00 (49.15%) |
| **Daily smoking** |  |
| No | 768.00 (86.78%) |
| Yes | 117.00 (13.22%) |
| **Physical activity** |  |
| High | 124.00 (14.01%) |
| Low | 587.00 (66.33%) |
| Moderate | 174.00 (19.66%) |
| **Body mass index** |  |
| Normal weight | 309.00 (34.92%) |
| Obesity | 225.00 (25.42%) |
| Overweight | 351.00 (39.66%) |
| **MetS - ATPIII** |  |
| No | 508.00 (57.40%) |
| Yes | 377.00 (42.60%) |
| **MetS - IDF** |  |
| No | 471.00 (53.22%) |
| Yes | 414.00 (46.78%) |
| **MetS - WHO** |  |
| No | 447.00 (50.51%) |
| Yes | 438.00 (49.49%) |
| **MetS - Total** |  |
| No | 294.00 (33.22%) |
| Yes | 591.00 (66.78%) |

Source: self made

In supplementary material 1, the bivariate table is presented, reflecting the percentage values in rows according to each probable associated factor. The graph of the percentage distribution of each component of MetS, according to its definition, is in supplementary material 2. In this, it is observed that the percentage levels could vary according to the cut-off point used in each component; it is much more distinct for abdominal waist according to IDF and ATPIII. Likewise, there were distinctions with fasting glucose alteration and elevated blood pressure, according to WHO.

Table 2 shows the multivariable regression analysis and the statistically significant associations between various factors and the presence of MetS, according to different diagnostic criteria.

**Table 2.** Multivariable regression analysis of the factors associated with MetS according to ATPIII, IDF, and WHO.

| Characteristic | Metabolic syndrome - ATPIII |  |  | Metabolic syndrome - IDF |  |  | Metabolic syndrome - WHO |  |  |
| --- | --- | --- | --- | --- | --- | --- | --- | --- | --- |
|  | aPR* | 95% CI | p-value | aPR* | 95% CI | p-value | aPR* | 95% CI | p-value |
| Sex |  |  |  |  |  |  |  |  |  |
| Female | — | — |  | — | — |  | — | — |  |
| Male | 0.70 | 0.56, 0.89 | <0.001 | 2.67 | 2.14, 3.33 | <0.001 | 1.25 | 1.02, 1.53 | <0.001 |
| Age group |  |  |  |  |  |  |  |  |  |
| 18 to 29 years old | — | — |  | — | — |  | — | — |  |
| 30 to 39 years old | 1.37 | 0.96, 1.99 | 0.014 | 1.35 | 0.97, 1.88 | 0.004 | 1.03 | 0.77, 1.39 | 0.739 |
| 40 to 49 years old | 1.60 | 1.12, 2.32 | <0.001 | 1.49 | 1.07, 2.10 | <0.001 | 1.05 | 0.77, 1.42 | 0.641 |
| 50 to 59 years old | 1.63 | 1.14, 2.37 | <0.001 | 1.51 | 1.08, 2.12 | <0.001 | 1.2 | 0.89, 1.62 | 0.075 |
| Civil status |  |  |  |  |  |  |  |  |  |
| Single | — | — |  | — | — |  | — | — |  |
| With couple | 1.2 | 0.94, 1.55 | 0.017 | 0.94 | 0.74, 1.19 | 0.38 | 1.01 | 0.81, 1.28 | 0.847 |
| Educational level |  |  |  |  |  |  |  |  |  |
| Higher | — | — |  | — | — |  | — | — |  |
| Secondary | 1.03 | 0.81, 1.31 | 0.673 | 1.07 | 0.85, 1.34 | 0.327 | 1.06 | 0.85, 1.31 | 0.447 |
| Until primary | 1.01 | 0.73, 1.40 | 0.91 | 1.51 | 1.09, 2.07 | <0.001 | 0.95 | 0.69, 1.29 | 0.613 |
| Natural region |  |  |  |  |  |  |  |  |  |
| Jungle | — | — | — | — | — | — | — | — | — |
| Metropolitan Lima | 0.8 | 0.58, 1.13 | **0.021** | 0.98 | 0.70, 1.39 | 0.871 | 0.8 | 0.60, 1.09 | **0.014** |
| Mountain Range | 0.66 | 0.43, 0.99 | **0.004** | 0.91 | 0.62, 1.33 | 0.481 | 0.36 | 0.24, 0.55 | **<0.001** |
| Rest of coast | 0.97 | 0.70, 1.37 | 0.777 | 1.15 | 0.82, 1.62 | 0.19 | 0.97 | 0.72, 1.32 | 0.756 |
| **Area of residence** |  |  |  |  |  |  |  |  |  |
| Rural | — | — | — | — | — | — | — | — | — |
| Urban | 0.98 | 0.72, 1.34 | 0.835 | 1.07 | 0.79, 1.46 | 0.494 | 1.12 | 0.83, 1.51 | 0.27 |
| **Wealth index** |  |  |  |  |  |  |  |  |  |
| No poor | — | — | — | — | — | — | — | — | — |
| Poor | 0.95 | 0.68, 1.31 | 0.631 | 0.95 | 0.69, 1.30 | 0.633 | 1.17 | 0.86, 1.58 | 0.122 |
| **Alcohol consumption** |  |  |  |  |  |  |  |  |  |
| No | — | — | — | — | — | — | — | — | — |
| Yes | 0.9 | 0.72, 1.13 | 0.122 | 1.11 | 0.90, 1.38 | 0.098 | 1.09 | 0.88, 1.34 | 0.22 |
| **Daily smoking** |  |  |  |  |  |  |  |  |  |
| No | — | — | — | — | — | — | — | — | — |
| Yes | 1.41 | 1.02, 1.92 | **<0.001** | 1.05 | 0.79, 1.38 | 0.534 | 1.17 | 0.88, 1.53 | 0.069 |
| **Physical activity** |  |  |  |  |  |  |  |  |  |
| High | — | — | — | — | — | — | — | — | — |
| Low | 1.14 | 0.84, 1.57 | 0.151 | 1.04 | 0.78, 1.40 | 0.713 | 1.02 | 0.78, 1.36 | 0.808 |
| Moderate | 1.28 | 0.90, 1.83 | **0.015** | 1.07 | 0.77, 1.51 | 0.525 | 1.09 | 0.79, 1.50 | 0.442 |
| **Body mass index** |  |  |  |  |  |  |  |  |  |
| Normal weight | — | — | — | — | — | — | — | — | — |
| Overweight | 4.27 | 2.88, 6.56 | **<0.001** | 2.52 | 1.87, 3.43 | **<0.001** | 1.35 | 1.05, 1.74 | **0.001** |
| Obesity | 7.18 | 4.81, 11.1 | **<0.001** | 4.10 | 3.00, 5.66 | **<0.001** | 1.74 | 1.33, 2.30 | **<0.001** |

\*Adjusted for sex, group age, year, educational level, civil status, natural region, area of residence, wealth index, daily smoking, alcohol consumption, physical activity and body mass index
aPR: Adjusted prevalence ratio. 95% CI: 95% confidence interval
Source: self made

According to the ATPIII criteria, males had a lower prevalence versus females (aPR: 0.7; 95% CI: 0.56, 0.89). The age groups of 30 to 39 years (aPR: 1.37; 95% CI: 0.96, 1.99), 40 to 49 years (aPR: 1.6; 95% CI: 1.12, 2.32) and 50 to 59 years (aPR: 1.63; 95% CI: 1.14, 2.37) showed a higher prevalence versus the group of 18 to 29 years. Those living as a couple also showed a higher prevalence compared to singles (aPR: 1.2; 95% CI: 0.94, 1.55). Individuals from the mountain region had a lower prevalence versus those from the jungle (aPR: 0.66; 95% CI: 0.43, 0.99). Daily smokers showed a higher prevalence (aPR: 1.41; 95% CI: 1.02, 1.92); those who practiced moderate physical activity, also a higher prevalence (aPR: 1.28; 95% CI: 0.90, 1.83) and those who were overweight (aPR: 4.27; 95% CI: 2.88, 6.56) and obese (aPR: 7.18; 95% CI: 4.81, 11.1), a higher prevalence versus those of normal weight.

According to the IDF criteria, men had a higher prevalence versus women (aPR: 2.67; 95% CI: 2.14, 3.33); the age groups of 30 to 39 years (aPR: 1.35; 95% CI: 0.97, 1.88), 40 to 49 years (aPR: 1.49; 95% CI: 1.07, 2.10) and 50 to 59 years (aPR: 1.51; 95% CI: 1.08, 2.12), higher prevalence versus the group of 18 to 29 years; those with up to primary education, a higher prevalence versus those with higher education (aPR: 1.51; 95% CI: 1.09, 2.07) and people with overweight (aPR: 2.52; 95% CI: 1.87, 3.43) and obesity (aPR: 4.1; 95% CI: 3.00, 5.66), a higher prevalence versus those of normal weight.

According to WHO, males had a higher prevalence versus females (aPR: 1.25; 95% CI: 1.02, 1.53); those from the mountain region, lower prevalence when compared with those from the jungle (aPR: 0.36; 95% CI: 0.24, 0.55). And those with overweight (aPR: 1.35; 95% CI: 1.05, 1.74) and obesity (aPR: 1.74; 95% CI: 1.33, 2.30), higher prevalence versus those of normal weight.

The agreement between the three criteria was evaluated through the Kappa index. The agreement between IDF with the WHO and ATPIII criteria was 0.42 (p<0.001) and 0.45 (p<0.001), respectively, while the agreement between ATPIII and the WHO criteria was 0.44 (p<0.001). On the other hand, in the Venn diagram, Figure 1, it can be seen that, in isolation, the ATPIII, IDF, and WHO criteria represented 7.4%, 10%, and 11.5% of the cases, respectively. However, when the intersections between the criteria were taken into account, higher proportions were appreciated. Specifically, the combination of the ATPIII and IDF criteria represented 8.5% of the cases; the combination of the IDF and WHO criteria, 14.7% of the cases, and the combination of the ATPIII and WHO criteria, 11.5% of the cases.

**Figure 1.**
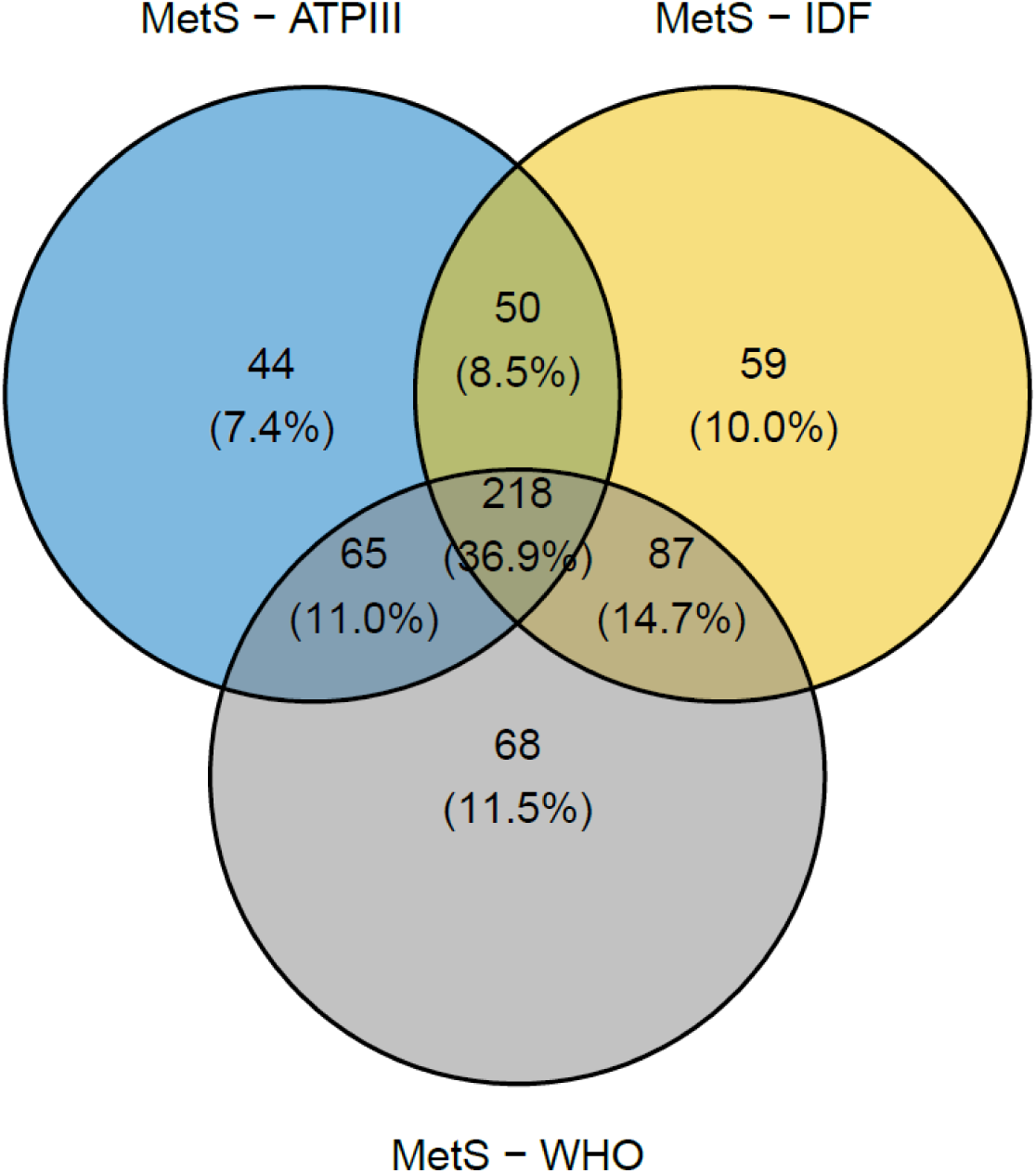
Venn diagram of the MetS according to ATPIII, WHO and IDF.

## DISCUSSION

### Prevalence of MetS according to the defined criterion

In our study, it was found that the prevalence of MetS in the Peruvian population was 49.49%, according to the WHO criteria; 42.60%, according to the ATPIII criterion, and 46.78%, according to the IDF criterion. That is, the prevalence of MetS varies according to the criterion used for its definition.

These findings are similar to those of others globally. A study was conducted in Iran found that the prevalence, according to the ATPIII criteria, was 29%, while, according to the IDF and WHO criteria, the total prevalence of MetS was 38% and 30%, respectively ^(12)^. In Mexico, on the other hand, a study determined that the prevalence of MetS, based on different criteria, was: IDF (54%), ATP III (36%) and WHO (31%) ^(11)^. Likewise, a study conducted in Sri Lanka in people with type 2 diabetes found that the prevalence of MetS was highest according to the WHO definition (70%), followed by the IDF (44%) and ATPIII (29%) ^(19)^. The study conducted in Mongolia by Myagmar-Ochir et al. (20), found that the prevalence of MetS in urban dwellers varied according to the definition used. The ATPIII, IDF, and JIS criteria yielded respective prevalence rates of 19.4%, 23.6%, and 25.4% according to this study’s findings on MetS.

While the three definitions of MetS each provide valuable insights, it is notable that their estimated prevalences vary substantially between nations, usefully illustrating intercountry disparities in the clustering of cardiometabolic risk factors. In our manuscript, the criterion according to the WHO was the highest, similar to the study in Sri Lanka ^(19)^, while in others, the highest was the IDF ^(11)^.

### Concordance of the three definitions of MetS

Regarding the concordance between the three definitions, there are studies that have found similar values and others discrepancies. In the work of Katulanda et al. ^(21)^, it was found that only about 25% of people have been recognized as having MetS simultaneously by the three definitions, suggesting that the three definitions identify a diverse group of people. This is true with the WHO definition, with almost 40% classified with MetS when the other two definitions did not. In contrast, most of the MetS recognized by the ATPIII definition have been identified with MetS by one of the other two definitions. In their work, the concordance of patients with MetS based on the IDF criteria with those of the WHO and ATPIII was 0.37 and 0.53, respectively, while it was 0.24 between ATPIII and WHO.

In the study by Myagmar-Ochir et al. ^(20)^ they compared the MetS definitions of the WHO, ATPIII, and IDF in an urban population of Mongolia. They found a moderate agreement between the ATPIII definitions and waist circumference and between the JIS definitions and fasting glucose and triglycerides. However, in the study by Subías-Perié et al., they found that the IDF definition and the JIS definition had the best agreement (k = 1.000). Meanwhile, the Finnish work by Haverinen et al.^(22)^ found that the IDF and JIS definitions had almost perfect agreement (k = 0.97), while the agreement between the ATPIII and JIS definitions was strong (k = 0.80).

It is important to highlight the importance of considering many definitions when studying the prevalence and concordance of MetS, since each one can capture different aspects of this complex condition and, therefore, could lead to different estimates. This variability in prevalence estimates, depending on the definition used, can have significant implications for public health planning and resource allocation.

### Factors associated with MetS

According to the ATPIII definition, males have a lower prevalence of MetS versus females, while according to the IDF and WHO definitions, males have a higher prevalence. The study’s discovery that being female independently connected to the existence of MetS, with a modified odds ratio of 4.69 ^(23)^, aligns with a recent investigation performed in Asmara, Eritrea, which also deduced a comparable relationship between the female sex and MetS using a parallel analysis. Nonetheless, given that the study was conducted exclusively among elderly individuals, it is imperative to underscore how the population’s advanced age could potentially impact the disparity seen in rates of MetS between genders addressed in the article. On the other hand, in the case of the IDF, this result is consistent with a recent work that examined the correlation of the siMS score, a quantification method for metabolic syndrome, with insulin resistance and other cofounding factors of metabolic syndrome in 451 obese patients with pre-metabolic syndrome and metabolic syndrome. In this research, the IDF classification was applied to diagnose metabolic syndrome, and it was found that males had a higher prevalence of MetS versus females ^(24)^.

The discrepancy in the association between sex and the prevalence of MetS, according to the ATPIII and IDF criteria, may be due to differences in the definitions of these. The ATPIII criterion necessitates meeting three or more of the subsequent factors must be met: abdominal obesity, elevated triglycerides, low HDL, high blood pressure, as well as elevated fasting glucose levels. On the other hand, the IDF definition places particular emphasis on central obesity (measured as waist circumference) along with the presence of two or more of the other factors. It is likely that the males in our sample had fewer metabolic risk factors, according to the ATPIII definition, plus a higher prevalence of central obesity, which would classify them as MetS according to the IDF definition ^(25,26)^.

The age ranges from thirty to fifty-nine exhibited a greater existence of MetS compared to the cluster from eighteen to twenty-nine, suggesting that the threat of MetS increases with age. This consequence agrees with previous reports showing the occurrence of MetS and the tendency to rise as time passes ^(23)^.

Mahmoud and Sulaiman found in 2022 that whether you’re married or not really mattered for your health. Being hitched, divorced/split up/gone was surely tied to a big risk of problems versus being lone ^(27)^. These discoveries suggest that if you’re married or not may play a part in how many folks have troubles, even if we ain’t totally clear on why and it’s down to lots of stuff. It’s like how your life changes when you get spliced, like what you eat and how much you move around, which could add to a higher chance of issues. Likewise, stress and changes to who’s there for you, which may come with leaving or being left, may also have an effect on how your body works ^(25)^.

Based on the standards of the ATPIII and WHO, it was found in our research that living in the highlands provided protection against metabolic syndrome as opposed to living in the jungle. This discovery aligns with the findings of a study performed in Brazil - urban living was connected with an increased risk of MetS versus rural living ^(28)^. Additionally, a report in China discovered that city living correlated to a higher chance of MetS. Likely, the differences in lifestyle, diet and physical activity between areas contribute to the variances in the chance of metabolic syndrome. However, further research remains necessary to investigate these potential rationales more thoroughly.

This report was looking at how smoking can influence MetS. They did a study in South Korea where they found that guys with over four unhealthy habits including cig smoking had a higher chance of dealing with MetS. It also said those with more bad lifestyle choices seemed to have more cases of MetS. Smoking could also be tied to high blood pressure, stomach fat, and high triglyceride levels too, which are big parts of MetS. This proves the idea that puffing would help cause MetS in a few ways, like assisting insulin resistance, long-term swelling, and dyslipidemia ^(30)^.

Regarding physical activity, in a study developed by Wewege et al ^(31)^. While our investigation determined that a moderate level of physical exertion correlated with an increased occurrence of MetS as defined by ATPIII standards, contradicting results have been found elsewhere, as one study discovered that physical activity of a moderate to vigorous nature associated with a lower prevalence of MetS, contrasting with our conclusions. The potential disparity could stem from divergences in how moderate physical exercise was characterized and the demographic examined. While regular physical activity undoubtedly represents an important element in both the avoidance and treatment of MetS, what must be stressed is its truly crucial role as a component for successfully preventing and managing this condition.

In our findings on the level of education, it was shown that those with education up to primary school showed a higher prevalence of MetS versus those with higher education according to the IDF criteria. The findings corroborate and complement what has already been released concerning this study area in a way that utilizes a more sophisticated syntactic construction. In a study conducted by Agyemang et al ^(32)^. The research findings demonstrated that individuals with less educational attainment confronted an elevated probability of experiencing Metabolic Syndrome. This potential explanation resides in the possibility that individuals with less attained education possess decreased health awareness and face impediments to accessing preventative healthcare, each of which could heighten their susceptibility to developing MetS. However, the research led by Ford and colleagues also found that ^(33)^, also found that the prevalence of MetS increased with the decrease in the level of education. These findings highlight the necessity of implementing public health initiatives focused on raising consciousness and understanding pertaining to MetS, specifically for individuals with more limited educational backgrounds.

On the other hand, BMI was strongly associated with MetS according to the three criteria used. Our findings demonstrated that the occurrence rates for MetS were determined to be greater amongst both individuals deemed as carrying excess weight and those judged as obese ^(34)^. Consistent with suggestions from prior reports, this discovery echoes that BMI represents a primary hazard contributing to the development of MetS. A study conducted in China by Li and colleagues discovered that body mass index was a noteworthy forecaster of metabolic syndrome, irrespective of the standards employed to characterize metabolic syndrome as referenced by the thirty-fourth source. Likewise, the research conducted by Zabetian and colleagues was comparable ^(35)^ found that weight reduction was a protective factor against MetS. The results corroborate the notion that an elevated BMI, signifying being overweight or obese, represents a significant threat to developing MetS.

Research into the differing metabolic functions of separate fat stores has illuminated that while body mass index offers a suitable measure of total adipose tissue amounts, it is unable to differentiate between subcutaneous and intra-abdominal deposits; intra-abdominal fat has demonstrated itself to be more metabolically active and more robustly linked to metabolic syndrome than subcutaneous fat supplies. While BMI provides insight into the likelihood of MetS, it alone does not fully account for the threat posed by where fat accumulates on the body.

### Study limitations

The cross-sectional nature of our manuscript restricts the ability to establish causal relationships. While various factors have been linked to the presence of MetS, the complex interplay between potential causes and outcomes precludes confirming with certainty whether these linkages represent causative mechanisms or resultant conditions. Also, although we have controlled for potentially confounding factors, the possibility of residual confusion cannot be completely eliminated, due to variables not measured or measured inaccurately. For instance, due to an inability to account for additional dietary variables like sleep quality or psychological conditions that may influence the frequency of MetS, we were incapable of adjusting for such extra factors impacting the prevalence of the metabolic syndrome.

Ultimately, while considering three diverse descriptions of MetS, each maintains particular constraints and may fail to comprise the full scope of MetS’s multifaceted nature completely. Although this research offers preliminary understandings, more rigorous exploration is merited so as to substantiate the outcomes owing to the inherent constraints and necessity for duplication in independent analyses.

### Conclusions

The prevalence of MetS in the studied population varies significantly depending on the diagnostic criteria used. Specifically, the prevalence of MetS was higher when the WHO definition was used versus the IDF and ATPIII definitions.

Various discerned factors regarding the propensity of MetS to manifest itself included one’s age and gender as well as marital status, smoking habits, place of residence, physical activity levels, and body mass index measurements. However, the association of these factors with MetS also varied depending on the definition used.

These results emphasize the importance of using a consistent and widely accepted definition of MetS in research and clinical practice. Furthermore, the findings imply that approaches for both preempting and addressing MetS ought to be tailored to the distinctive qualities of the community in question, while also considering the diverse array of potential contributing aspects.

Further investigations are recommended to confirm our findings and further explore the differences in the prevalence and associated factors of MetS according to different definitions. In addition, efforts should be made to harmonize the definitions of MetS and develop clear, evidence-based guidelines for its diagnosis and management.

## Acknowledgments

A special thanks to the members of Instituto de Investigación en Ciencias Biomédicas de la Universidad Ricardo Palma who provided valuable comments during the preparation of this study.

## Financial disclosure

This study is self-financed.

## Conflict of interest

The authors declare no conflict of interest.

## Informed consent

The informed consent was obtained.

## Author contributions

Victor Juan Vera-Ponce, Fiorella E. Zuzunaga-Montoya, Luisa Erika Milagros Vásquez Romero and Joan A. Loayza-Castro participated in the genesis of the idea and project design. Fiorella E. Zuzunaga-Montoya, Luisa Erika Milagros Vásquez, and Mario J. Valladares-Garrido oversaw the data collection, interpretation, and analysis of results. Joan A. Loayza-Castro and Victor Juan Vera-Ponce contributed to the preparation of the manuscript of this research paper.

## Data availability

The data supporting the findings of this study can be accessed through the following link: https://www.datosabiertos.gob.pe/dataset/estado-nutricional-en-adultos-de-18-59-a%C3%B1os-per%C3%BA-2017-%E2%80%93-2018

## Supplementary material 1

**Table.**
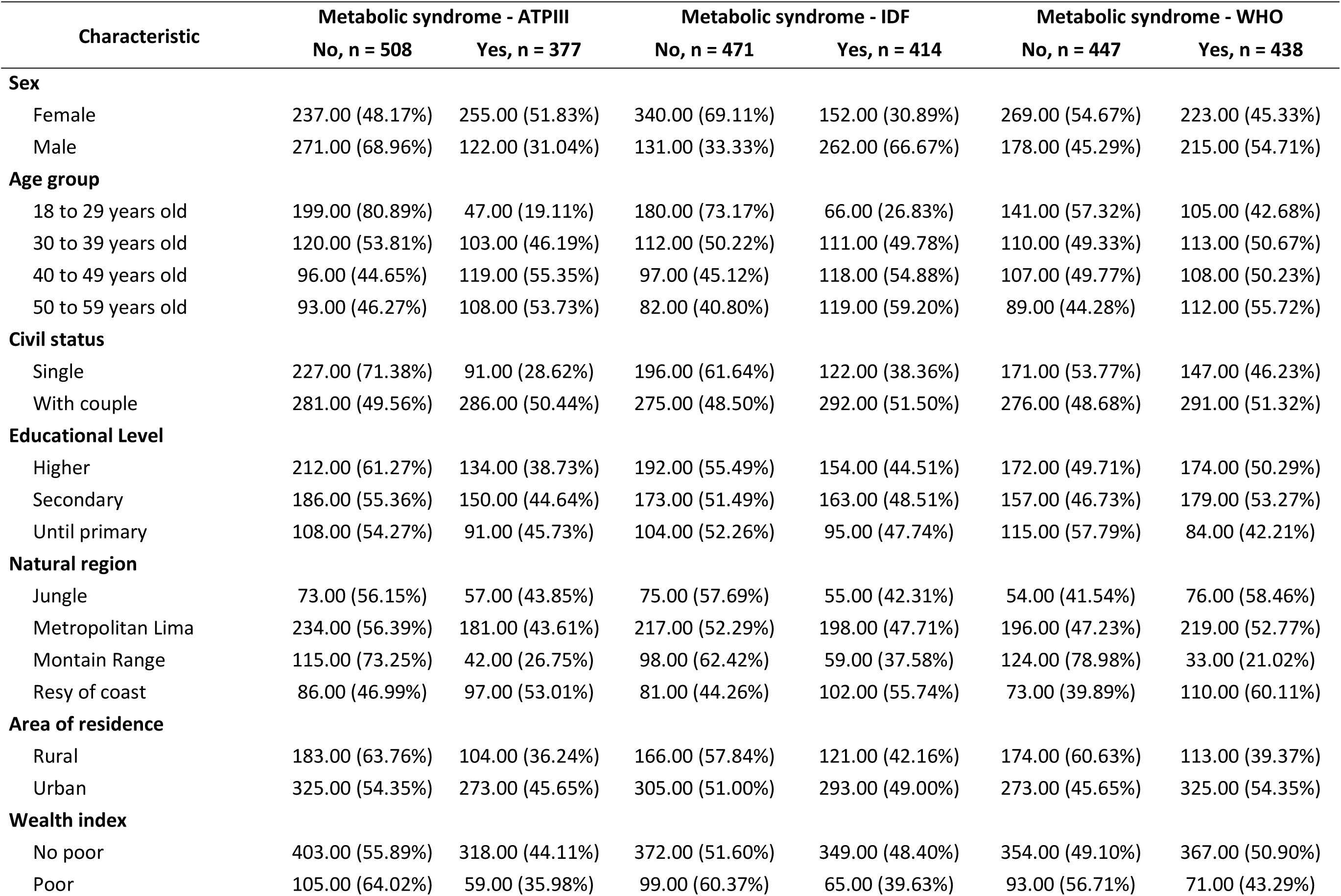
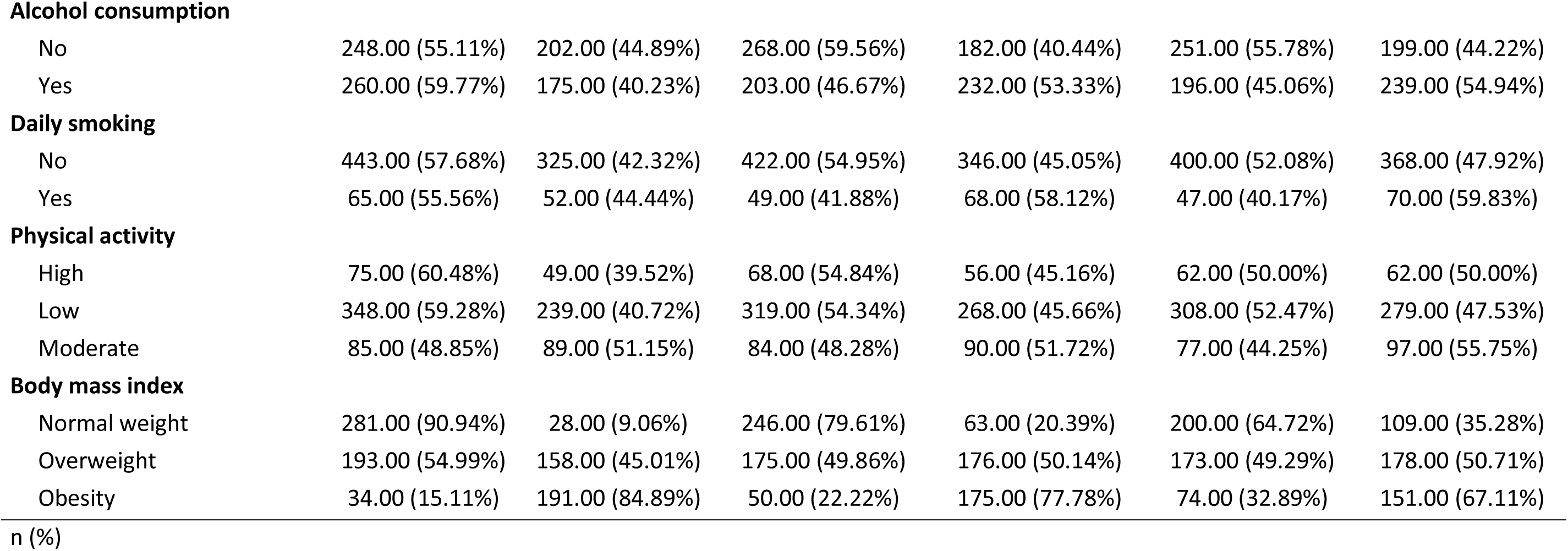
Bivariate analysis of factors associated with MetS according to ATPIII, IDF and WHO.

| Characteristic | Metabolic syndrome - ATPIII |  | Metabolic syndrome - IDF |  | Metabolic syndrome - WHO |  |
| --- | --- | --- | --- | --- | --- | --- |
|  | No, n = 508 | Yes, n = 377 | No, n = 471 | Yes, n = 414 | No, n = 447 | Yes, n = 438 |
| **Sex** |  |  |  |  |  |  |
| Female | 237.00 (48.17%) | 255.00 (51.83%) | 340.00 (69.11%) | 152.00 (30.89%) | 269.00 (54.67%) | 223.00 (45.33%) |
| Male | 271.00 (68.96%) | 122.00 (31.04%) | 131.00 (33.33%) | 262.00 (66.67%) | 178.00 (45.29%) | 215.00 (54.71%) |
| **Age group** |  |  |  |  |  |  |
| 18 to 29 years old | 199.00 (80.89%) | 47.00 (19.11%) | 180.00 (73.17%) | 66.00 (26.83%) | 141.00 (57.32%) | 105.00 (42.68%) |
| 30 to 39 years old | 120.00 (53.81%) | 103.00 (46.19%) | 112.00 (50.22%) | 111.00 (49.78%) | 110.00 (49.33%) | 113.00 (50.67%) |
| 40 to 49 years old | 96.00 (44.65%) | 119.00 (55.35%) | 97.00 (45.12%) | 118.00 (54.88%) | 107.00 (49.77%) | 108.00 (50.23%) |
| 50 to 59 years old | 93.00 (46.27%) | 108.00 (53.73%) | 82.00 (40.80%) | 119.00 (59.20%) | 89.00 (44.28%) | 112.00 (55.72%) |
| **Civil status** |  |  |  |  |  |  |
| Single | 227.00 (71.38%) | 91.00 (28.62%) | 196.00 (61.64%) | 122.00 (38.36%) | 171.00 (53.77%) | 147.00 (46.23%) |
| With couple | 281.00 (49.56%) | 286.00 (50.44%) | 275.00 (48.50%) | 292.00 (51.50%) | 276.00 (48.68%) | 291.00 (51.32%) |
| **Educational Level** |  |  |  |  |  |  |
| Higher | 212.00 (61.27%) | 134.00 (38.73%) | 192.00 (55.49%) | 154.00 (44.51%) | 172.00 (49.71%) | 174.00 (50.29%) |
| Secondary | 186.00 (55.36%) | 150.00 (44.64%) | 173.00 (51.49%) | 163.00 (48.51%) | 157.00 (46.73%) | 179.00 (53.27%) |
| Until primary | 108.00 (54.27%) | 91.00 (45.73%) | 104.00 (52.26%) | 95.00 (47.74%) | 115.00 (57.79%) | 84.00 (42.21%) |
| **Natural region** |  |  |  |  |  |  |
| Jungle | 73.00 (56.15%) | 57.00 (43.85%) | 75.00 (57.69%) | 55.00 (42.31%) | 54.00 (41.54%) | 76.00 (58.46%) |
| Metropolitan Lima | 234.00 (56.39%) | 181.00 (43.61%) | 217.00 (52.29%) | 198.00 (47.71%) | 196.00 (47.23%) | 219.00 (52.77%) |
| Mountain Range | 115.00 (73.25%) | 42.00 (26.75%) | 98.00 (62.42%) | 59.00 (37.58%) | 124.00 (78.98%) | 33.00 (21.02%) |
| Resy of coast | 86.00 (46.99%) | 97.00 (53.01%) | 81.00 (44.26%) | 102.00 (55.74%) | 73.00 (39.89%) | 110.00 (60.11%) |
| **Area of residence** |  |  |  |  |  |  |
| Rural | 183.00 (63.76%) | 104.00 (36.24%) | 166.00 (57.84%) | 121.00 (42.16%) | 174.00 (60.63%) | 113.00 (39.37%) |
| Urban | 325.00 (54.35%) | 273.00 (45.65%) | 305.00 (51.00%) | 293.00 (49.00%) | 273.00 (45.65%) | 325.00 (54.35%) |
| **Wealth index** |  |  |  |  |  |  |
| No poor | 403.00 (55.89%) | 318.00 (44.11%) | 372.00 (51.60%) | 349.00 (48.40%) | 354.00 (49.10%) | 367.00 (50.90%) |
| Poor | 105.00 (64.02%) | 59.00 (35.98%) | 99.00 (60.37%) | 65.00 (39.63%) | 93.00 (56.71%) | 71.00 (43.29%) |
| **Alcohol consumption** |  |  |  |  |  |  |
| No | 248.00 (55.11%) | 202.00 (44.89%) | 268.00 (59.56%) | 182.00 (40.44%) | 251.00 (55.78%) | 199.00 (44.22%) |
| Yes | 260.00 (59.77%) | 175.00 (40.23%) | 203.00 (46.67%) | 232.00 (53.33%) | 196.00 (45.06%) | 239.00 (54.94%) |
| **Daily smoking** |  |  |  |  |  |  |
| No | 443.00 (57.68%) | 325.00 (42.32%) | 422.00 (54.95%) | 346.00 (45.05%) | 400.00 (52.08%) | 368.00 (47.92%) |
| Yes | 65.00 (55.56%) | 52.00 (44.44%) | 49.00 (41.88%) | 68.00 (58.12%) | 47.00 (40.17%) | 70.00 (59.83%) |
| **Physical activity** |  |  |  |  |  |  |
| High | 75.00 (60.48%) | 49.00 (39.52%) | 68.00 (54.84%) | 56.00 (45.16%) | 62.00 (50.00%) | 62.00 (50.00%) |
| Low | 348.00 (59.28%) | 239.00 (40.72%) | 319.00 (54.34%) | 268.00 (45.66%) | 308.00 (52.47%) | 279.00 (47.53%) |
| Moderate | 85.00 (48.85%) | 89.00 (51.15%) | 84.00 (48.28%) | 90.00 (51.72%) | 77.00 (44.25%) | 97.00 (55.75%) |
| **Body mass index** |  |  |  |  |  |  |
| Normal weight | 281.00 (90.94%) | 28.00 (9.06%) | 246.00 (79.61%) | 63.00 (20.39%) | 200.00 (64.72%) | 109.00 (35.28%) |
| Overweight | 193.00 (54.99%) | 158.00 (45.01%) | 175.00 (49.86%) | 176.00 (50.14%) | 173.00 (49.29%) | 178.00 (50.71%) |
| Obesity | 34.00 (15.11%) | 191.00 (84.89%) | 50.00 (22.22%) | 175.00 (77.78%) | 74.00 (32.89%) | 151.00 (67.11%) |

## Supplementary material 2

**Figure 2.**
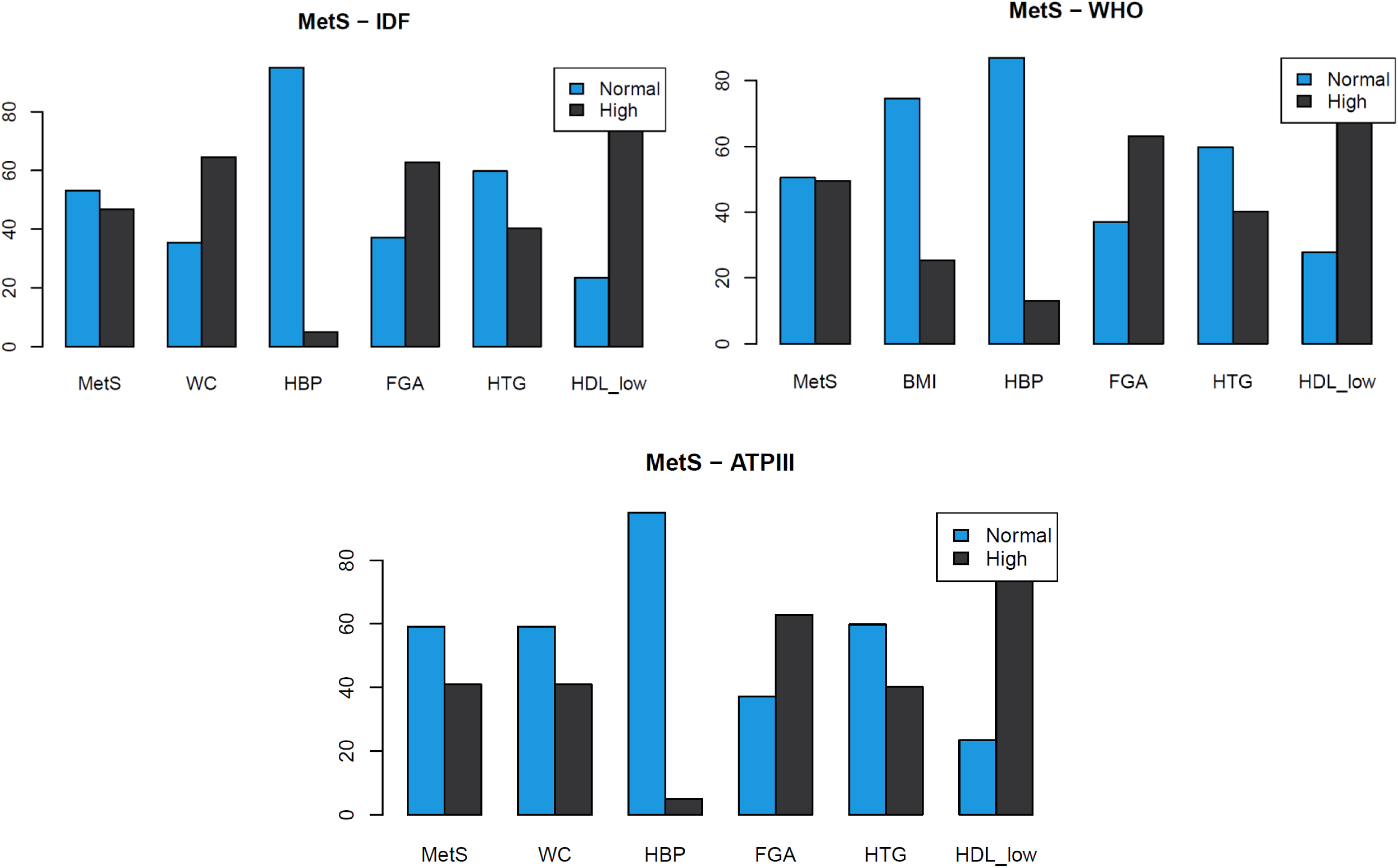
Prevalence of MetS components according to: a) IDF; b) WHO, c) ATPII

## Notes

### Competing Interest Statement

The authors have declared no competing interest.

### Author Declarations

The study used freely available data and in the public domain online, that were originally located at: http://iinei.inei.gob.pe/microdatos/

## Bibliographic references

1. Shalitin S, Giannini C. Obesity, Metabolic Syndrome, and Nutrition. World Rev Nutr Diet. 2023;126:47–69. doi:10.1159/000527938

2. Furuta K, Tang X, Islam S, Tapia A, Chen ZB, Ibrahim SH. Endotheliopathy in the metabolic syndrome: Mechanisms and clinical implications. Pharmacol Ther. 2023;244:108372. doi:10.1016/j.pharmthera.2023.108372

3. Mahadevan M, Bose M, Gawron KM, Blumberg R. Metabolic Syndrome and Chronic Disease Risk in South Asian Immigrants: A Review of Prevalence, Factors, and Interventions. Healthcare. 2023;11(5):720. doi:10.3390/healthcare11050720

4. Hirode G, Wong RJ. Trends in the Prevalence of Metabolic Syndrome in the United States, 2011-2016. JAMA. 2020;323(24):2526–8. doi:10.1001/jama.2020.4501

5. Zhang R, Sun J, Wang C, Wang X, Zhao P, Yuan Y, et al. The Racial Disparities in the Epidemic of Metabolic Syndrome With Increased Age: A Study From 28,049 Chinese and American Adults. Front Public Health. 2022;9:797183. doi:10.3389/fpubh.2021.797183

6. Márquez-Sandoval F, Macedo-Ojeda G, Viramontes-Hörner D, Fernández Ballart JD, Salas Salvadó J, Vizmanos B. The prevalence of metabolic syndrome in Latin America: a systematic review. Public Health Nutr. 2011;14(10):1702–13. doi:10.1017/S1368980010003320

7. Arbañil-Huamán HC. Síndrome metabólico: Definición y prevalencia. Revista Peruana de Ginecología y Obstetricia. 2011;57(4):233–6.

8. Adams KJ, Chirinos JL. Prevalencia de factores de riesgo para síndrome metabólico y sus componentes en usuarios de comedores populares en un distrito de Lima, Perú. Revista Peruana de Medicina Experimental y Salud Pública. 2018;35(1):39–45. doi:10.17843/rpmesp.2018.351.3598

9. Bernabé-Ortiz A, Carrillo-Larco RM, Gilman RH, Checkley W, Smeeth L, Miranda JJ, et al. Contribution of modifiable risk factors for hypertension and type-2 diabetes in Peruvian resource-limited settings. J Epidemiol Community Health. 2016;70(1):49–55. doi:10.1136/jech-2015-205988

10. Ahima RS, editor. Metabolic Syndrome: A Comprehensive Textbook [Internet]. Cham: Springer International Publishing; 2014 [citado el 28 de julio de 2023]. doi:10.1007/978-3-319-12125-3

11. Gutiérrez-Solis AL, Datta Banik S, Méndez-González RM. Prevalence of Metabolic Syndrome in Mexico: A Systematic Review and Meta-Analysis. Metab Syndr Relat Disord. 2018;16(8):395–405. doi:10.1089/met.2017.0157

12. Dalvand S, Niksima SH, Meshkani R, Ghanei Gheshlagh R, Sadegh-Nejadi S, Kooti W, et al. Prevalence of Metabolic Syndrome among Iranian Population: A Systematic Review and Meta-analysis. Iran J Public Health. 2017;46(4):456–67.

13. Athyros VG, Ganotakis ES, Elisaf MS, Liberopoulos EN, Goudevenos IA, Karagiannis A, et al. Prevalence of vascular disease in metabolic syndrome using three proposed definitions. Int J Cardiol. 2007;117(2):204–10. doi:10.1016/j.ijcard.2006.04.078

14. Centro Nacional de Alimentación y Nutrición. Estado Nutricional En Adultos de 18 a 59 Años VIANEV 2017–2018. Instituto Nacional de Salud: Lima, Peru. 2021;191.

15. von Elm E, G. Altman D, Egger M, J. Pocock S, C. Gotzsche P, P. Vandenbroucke J. Declaración de la Iniciativa STROBE (Strengthening the Reporting of Observational studies in Epidemiology): directrices para la comunicación de estudios observacionales. 2008;22(2):144–50. doi:https://www.equator-network.org/wp-content/uploads/2015/10/STROBE_Spanish.pdf

16. Blackburn P, Lemieux I, Alméras N, Bergeron J, Côté M, Tremblay A, et al. The hypertriglyceridemic waist phenotype versus the National Cholesterol Education Program-Adult Treatment Panel III and International Diabetes Federation clinical criteria to identify high-risk men with an altered cardiometabolic risk profile. Metab Clin Exp. 2009;58(8):1123–30. doi:10.1016/j.metabol.2009.03.012

17. Alberti KGMM, Zimmet P, Shaw J. Metabolic syndrome--a new world-wide definition. A Consensus Statement from the International Diabetes Federation. Diabet Med. 2006;23(5):469–80. doi:10.1111/j.1464-5491.2006.01858.x

18. Definition, diagnosis and classification of diabetes mellitus and its complications : report of a WHO consultation. Part 1, Diagnosis and classification of diabetes mellitus [Internet]. [citado el 28 de julio de 2023]. Disponible en: https://apps.who.int/iris/handle/10665/66040

19. Herath HMM, Weerasinghe NP, Weerarathna TP, Amarathunga A. A Comparison of the Prevalence of the Metabolic Syndrome among Sri Lankan Patients with Type 2 Diabetes Mellitus Using WHO, NCEP-ATP III, and IDF Definitions. Int J Chronic Dis. 2018;2018:7813537. doi:10.1155/2018/7813537

20. Myagmar-Ochir E, Haruyama Y, Takaoka N, Takahashi K, Dashdorj N, Dashtseren M, et al. Comparison of Three Diagnostic Definitions of Metabolic Syndrome and Estimation of Its Prevalence in Mongolia. International Journal of Environmental Research and Public Health. 2023;20(6):4956. doi:10.3390/ijerph20064956

21. Katulanda P, Ranasinghe P, Jayawardana R, Sheriff R, Matthews DR. Metabolic syndrome among Sri Lankan adults: prevalence, patterns and correlates. Diabetol Metab Syndr. 2012;4(1):24. doi:10.1186/1758-5996-4-24

22. Haverinen E, Paalanen L, Palmieri L, Padron-Monedero A, Noguer-Zambrano I, Sarmiento Suárez R, et al. Comparison of metabolic syndrome prevalence using four different definitions –a population-based study in Finland. Archives of Public Health. 2021;79(1):231. doi:10.1186/s13690-021-00749-3

23. Achila OO, Araya M, Berhe AB, Haile NH, Tsige LK, Shifare BY, et al. Metabolic syndrome, associated factors and optimal waist circumference cut points: findings from a cross-sectional community-based study in the elderly population in Asmara, Eritrea. BMJ Open. 2022;12(2):e052296. doi:10.1136/bmjopen-2021-052296

24. Dimitrijevic-Sreckovic V, Petrovic H, Dobrosavljevic D, Colak E, Ivanovic N, Gostiljac D, et al. siMS score-method for quantification of metabolic syndrome, confirms co-founding factors of metabolic syndrome. Frontiers in Genetics [Internet]. 2023 [citado el 28 de julio de 2023];13. Disponible en: https://www.frontiersin.org/articles/10.3389/fgene.2022.1041383

25. Ye Y, Zhou Q, Dai W, Peng H, Zhou S, Tian H, et al. Gender differences in metabolic syndrome and its components in southern china using a healthy lifestyle index: a cross-sectional study. BMC Public Health. 2023;23(1):686. doi:10.1186/s12889-023-15584-0

26. Ezeani A, Tcheugui JBE, Agurs-Collins T. Sex/gender differences in metabolic syndrome among cancer survivors in the US: an NHANES analysis. J Cancer Surviv [Internet]. 2023 [citado el 28 de julio de 2023]; doi:10.1007/s11764-023-01404-2

27. Mahmoud I, Sulaiman N. Prevalence of Metabolic Syndrome and Associated Risk Factors in the United Arab Emirates: A Cross-Sectional Population-Based Study. Frontiers in Public Health [Internet]. 2022 [citado el 28 de julio de 2023];9. Disponible en: https://www.frontiersin.org/articles/10.3389/fpubh.2021.811006

28. de Siqueira Valadares LT, de Souza LSB, Salgado Júnior VA, de Freitas Bonomo L, de Macedo LR, Silva M. Prevalence of metabolic syndrome in Brazilian adults in the last 10 years: a systematic review and meta-analysis. BMC Public Health. 2022;22(1):327. doi:10.1186/s12889-022-12753-5

29. Li R, Li W, Lun Z, Zhang H, Sun Z, Kanu JS, et al. Prevalence of metabolic syndrome in Mainland China: a meta-analysis of published studies. BMC Public Health. 2016;16:296. doi:10.1186/s12889-016-2870-y

30. Park YS, Kang SH, Jang S-I, Park E-C. Association between lifestyle factors and the risk of metabolic syndrome in the South Korea. Sci Rep. 2022;12(1):13356. doi:10.1038/s41598-022-17361-2

31. Wewege M, van den Berg R, Ward RE, Keech A. The effects of high-intensity interval training vs. moderate-intensity continuous training on body composition in overweight and obese adults: a systematic review and meta-analysis. Obes Rev. 2017;18(6):635–46. doi:10.1111/obr.12532

32. Agyemang C, Meeks K, Beune E, Owusu-Dabo E, Mockenhaupt FP, Addo J, et al. Obesity and type 2 diabetes in sub-Saharan Africans – Is the burden in today’s Africa similar to African migrants in Europe? The RODAM study. BMC Medicine. 2016;14(1):166. doi:10.1186/s12916-016-0709-0

33. Ford ES, Giles WH, Dietz WH. Prevalence of the metabolic syndrome among US adults: findings from the third National Health and Nutrition Examination Survey. JAMA. 2002;287(3):356–9. doi:10.1001/jama.287.3.356

34. Li S, Xiao J, Ji L, Weng J, Jia W, Lu J, et al. BMI and waist circumference are associated with impaired glucose metabolism and type 2 diabetes in normal weight Chinese adults. J Diabetes Complications. 2014;28(4):470–6. doi:10.1016/j.jdiacomp.2014.03.015

35. Zabetian A, Hadaegh F, Sarbakhsh P, Azizi F. Weight change and incident metabolic syndrome in Iranian men and women; a 3 year follow-up study. BMC Public Health. 2009;9:138. doi:10.1186/1471-2458-9-138

36. Neeland IJ, Ross R, Després J-P, Matsuzawa Y, Yamashita S, Shai I, et al. Visceral and ectopic fat, atherosclerosis, and cardiometabolic disease: a position statement. Lancet Diabetes Endocrinol. 2019;7(9):715–25. doi:10.1016/S2213-8587(19)30084-1

